# Remote effects of temporal lobe epilepsy surgery: long-term morphological changes after surgical resection

**DOI:** 10.1101/2020.11.05.20224873

**Authors:** T. Campbell Arnold, Lohith G. Kini, John M. Bernabei, Andrew Y. Revell, Sandhitsu R. Das, Joel M. Stein, Tim H. Lucas, Dario J. Englot, Victoria L. Morgan, Brian Litt, Kathryn A. Davis

**Author notes:** Corresponding author name: T. Campbell Arnold, Corresponding authors, Corresponding, Corresponding, Corresponding author address: Department of Bioengineering, University of Pennsylvania, 240 Skirkanich Hall, 210 S 33^rd^ St, Philadelphia PA 19104.

## Abstract

**Objective:** We present a semi-automated method for quantifying structural changes after epilepsy surgery that accounts for tissue deformation caused by resection. We demonstrate its utility by comparing the remote structural effects of two surgical approaches, the anterior temporal lobectomy (ATL) and the selective amygdalohippocampectomy (SAH).

**Methods:** We studied 37 temporal lobe epilepsy (TLE) patients who underwent resective surgery. Patients were treated with either an anterior temporal lobectomy (ATL, N=21) or a selective amygdalohippocampectomy (SAH, N=16). All patients received same-scanner MR imaging preoperatively and postoperatively (5+ months after surgery). To analyze structural changes in remote brain regions, we (1) implemented an automated method for segmenting resections with manual review, (2) applied cost function masking to the resection zone, and (3) estimated longitudinal cortical thickness changes using Advanced Normalization Tools (ANTs). We then compared post-operative changes in cortical thickness between the two surgical groups in brain regions outside the resected area.

**Results:** Patients treated with ATL exhibited significantly greater cortical thinning globally when compared to patients treated with SAH (*p* = 0.049). There were significant focal differences between the two treatment groups in the ipsilateral frontal lobe (superior medial and medial orbital regions) and insula (*p* > 0.001, *α* = 0.05 Bonferroni corrected). No significant effects were seen in the contralateral hemisphere.

**Significance:** We present and share a semi-automated pipeline for quantifying remote longitudinal changes in cortical thickness after neurosurgery. The technique is applicable to a broad array of applications, including surgical planning and mapping neuropsychological function to brain structure. Using this tool, we demonstrate that patients treated with SAH for refractory temporal lobe epilepsy have less postoperative cortical thinning in remote brain regions than those treated with ATL. We share all algorithm code and results to accelerate collaboration and clinical translation of our work.

**KEY POINTS BOX:**

- Different epilepsy surgical approaches lead to distinct patterns of postoperative cortical atrophy in remote brain regions
- Patients treated with SAH have less postoperative cortical thinning than patients treated with ATL
- The insula and frontal lobe demonstrated the greatest focal differences in postoperative cortical thinning when comparing SAH and ATL
- Postoperative cortical thinning analyses may inform surgical planning and our understanding of cognitive sequelae

## INTRODUCTION

Epilepsy affects sixty-five million people worldwide, with one third of patients suffering from drug-resistant epilepsy (DRE)^1,2^. Individuals suffering from DRE bear greater risk of premature death, injury and worsening quality of life, including psychosocial dysfunction^3^. In patients where a focal seizure onset zone can be localized, surgical resection has been established as an effective treatment for DRE^4^. The temporal lobe is the most common site of localization, with temporal lobe epilepsy (TLE) accounting for approximately ∼65% of DRE. While anterior temporal lobectomy is the most common surgical approach for patients with TLE, in recent years surgical options have expanded and now include more selective resections^5^, focal laser ablations^6^, and targeted intracranial neural stimulation^7^. These less invasive surgical approaches seek to achieve seizure control while preserving cognitive function and reducing surgical comorbidities^8,9^. However, comparisons of cognitive outcomes and seizure freedom between traditional resection and selective amygdalohippocampectomy have been mixed, which likely reflects a complex trade-off between preserving functional brain tissue and removing epileptogenic regions ^10–13^.

Across institutions, numerous surgical procedures are performed for epilepsy management, including anterior temporal lobectomy (ATL), selective amygdalohippocampectomy (SAH), lesionectomy, and focal ablation. Substantial evidence, including a rigorous randomized controlled trial and multi-institution meta-analyses, indicate that ATL is superior to continued antiepileptic medication in up to 70% of cases^4,14^. However, evidence indicates that more focal surgery, such as laser ablations, may lead to better cognitive outcomes^6,13^. Despite the growing list of surgical options, clinicians lack quantitative methods for assessing the impact of different procedures on brain structure and function. It is known that ongoing seizures are associated with accelerated atrophy throughout the brain, which contributes to cognitive decline^15^. However, comparatively little is known about how surgical procedures affect long-term brain morphometry. Quantitative neuroimaging could provide key insights into differences in long-term seizure-freedom and cognitive outcome between surgical treatments thus providing clinical guidance and improve patient outcomes^16^.

Surgical resection has remote consequences outside of local tissues that are removed, such as white matter atrophy^17^, however these remote changes are not well understood. While many studies delineate the effects of ongoing seizure on cortical thickness^18^, few quantify the change seen between presurgical and postsurgical imaging^19,20^. It is important to characterize downstream effects of surgical procedures as they may negatively affect patient outcome and lead to decreased rates of seizure freedom or increased cognitive side-effects from surgery. Furthermore, given a planned resection by the clinical team, quantitative measurements in combination with brain connectivity analyses could be used to model the remote effects of a proposed surgery on the brain^21,22^. Recent neurosurgical advances have refined resective therapies for DRE, but there remains a great need to better understand the effects of surgery on the postsurgical brain to guide future therapy.

In this study, we use advanced computational imaging tools to quantify changes throughout the brain and hypothesize that there are measurable changes in cortical thickness caused by epilepsy surgery. We propose that these changes are a function of the surgical technique employed. To understand the effects of surgical approach on brain structures, we developed quantitative techniques to evaluate postoperative imaging for cortical thickness changes in adjacent and remote brain regions. We quantify downstream effects of two common surgical procedures, the ATL and the SAH in 37 patients who underwent temporal lobe epilepsy surgery. We developed a semi-automated method for segmenting resection cavities and considering the extent of the resection zone when computing cortical thinning in the rest of the brain. We publicly share our code to allow others to validate and improve upon our methods for assessing post-operative changes in cortical thickness to accelerate translation into clinical practice.

## METHODS

### Patients

Thirty-seven patients who underwent surgery for localization-related epilepsy across two institutions, Hospital of the University of Pennsylvania (HUP, N=14) and Vanderbilt University Medical Center (VUMC, N=23), were recruited for this study. The protocol was approved by the Vanderbilt University Institutional Review Board and the Institutional Review Board of the University of Pennsylvania. All participants gave informed consent. Patients were age-matched across institution (HUP: 38.9 ± 13.3, VUMC: 39.5 ± 12.7) and were treated with either an ATL(N=21) or SAH (N=16). One patient treated with a focal laser ablation of the amygdala and hippocampus was included in the SAH group. Patient demographics and clinical characteristics are listed in **Table 1**.

**Table 1 –.**
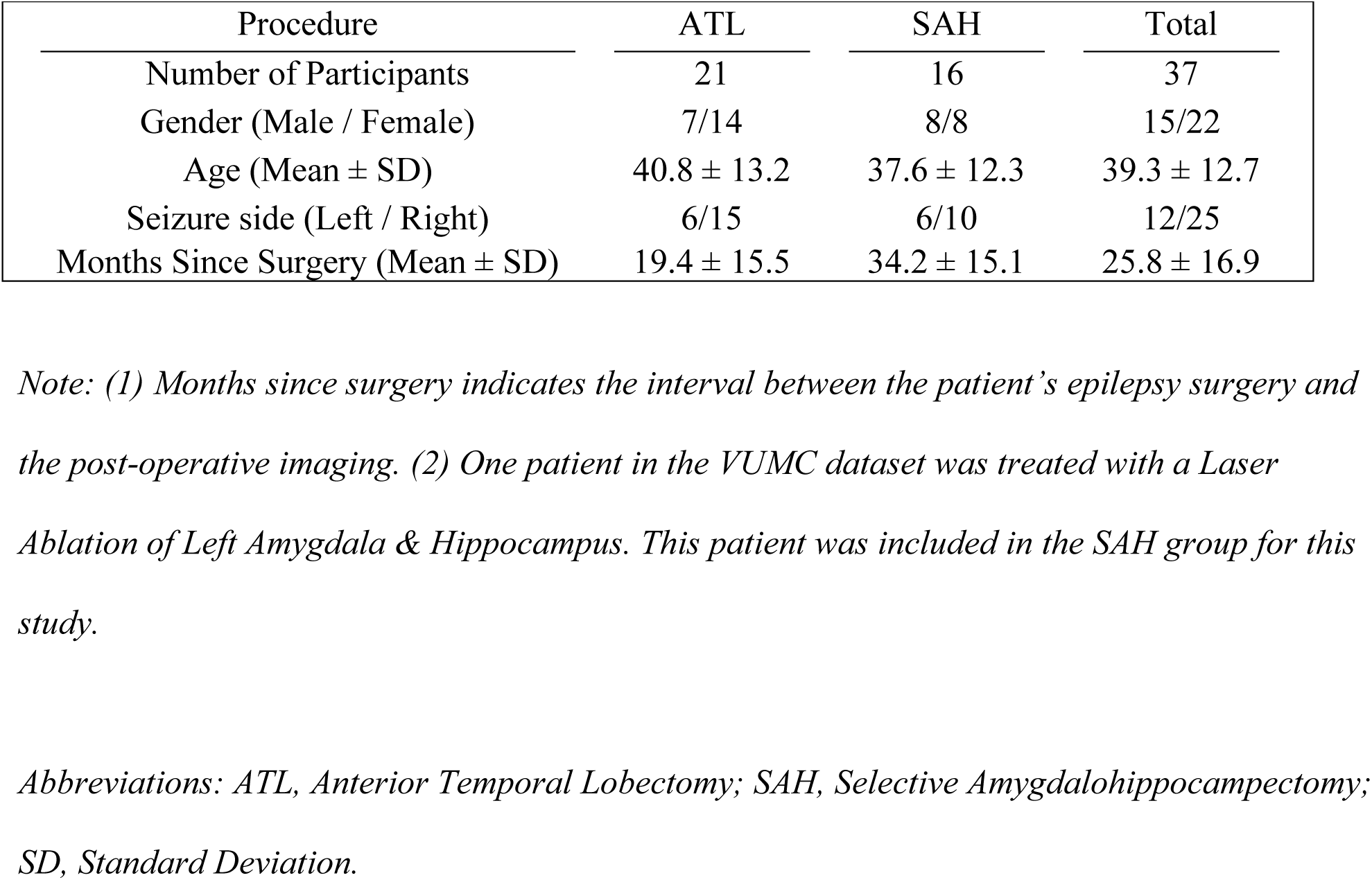
Demographics of patients included in study.

### Image Acquisition

All patients underwent a clinical epilepsy neuroimaging protocol both preoperatively and postoperatively (<5 months after surgery) as part of their standard care. The post-resection imaging protocol was acquired, on average, 26 months after implant and resection. 1 mm isotropic pre-implant T1-weighted MRI (T1w) and post-resection T1w images were acquired. Preoperative and postoperative images were collected on the same scanner to avoid the confounds of inter-scanner variability. Patients at HUP were imaged using a Siemens Trio scanner with a 64-channel head coil and received a T1w with the following acquisition parameters (0.98×0.98×1mm, TE=3.87ms, TR=1.62s, flip-angle=15). Patients at VUMC were imaged using a Phillips 3T scanner with a 32-channel head coil and received a T1w acquired with the following acquisition parameters (1mm isotropic, TE=4.61ms, TR=8.9ms, flip-angle= 8).

### Resection zone segmentation

Preoperative and postoperative images were post-processed using Advanced Normalization Tools (ANTs), an open-source neuroimaging software suite^23^. Preoperative images were registered to the postoperative image and the Atropos algorithm^24^ was used to segment images into six tissue types: cerebral spinal fluid (CSF), grey matter (GM), white matter (WM), deep grey matter (DGM), cerebellum, and brainstem. In preoperative images, the resection area is labeled as GM or WM, however after resection the same voxels are classified as CSF. To generate an automated estimation of the resection zone, the intersection between preoperative tissue segmentations (GM and WM) and the difference in pre-to-postoperative CSF segmentation was obtained^25^. Prior to subtraction, a gaussian smoothing kernel (sigma = 2) was passed over the probability maps to avoid large contrast due to alignment^26^. The extent of the resection mask can be modified by applying more stringent or liberal thresholds to tissue probability maps before calculating their intersection. In an ad hoc analysis, preoperative tissue classified as GM or WM with a probability greater than 50% and a pre-minus-post CSF difference greater than 25% yielded accurate localizations. To eliminate stray voxels, only the largest contiguous cluster of voxels in the mask was retained. Resection zones estimations were manually reviewed using the ITK-SNAP toolkit^27^ and necessary corrections were made manually by the authors.

### Cortical thinning estimation

We assessed cortical thickness using an adaptation of the well validated ANTs longitudinal cortical thickness pipeline^23,28^. For patients with cortical resections, the standard ANTs pipeline causes significant image distortion during deformable registration of postoperative images to a template image (**Figure 1**)^29^. This distortion results from WM and GM in the postoperative image erroneously being pulled into the resection cavity to better match the template image intensity values, ultimately leading to downstream inaccuracies in cortical thickness estimation. Cost function masking (CFM) has proven to be an effective method for preventing image distortion during registration in lesional brains^30^. In CFM, the abnormal region is segmented and excluded when performing subsequent registration calculations, thus preventing that region from influencing registration. To mitigate these errors, we adjusted the ANTs pipeline to include cost function masking of the resection zone (**Figure 2**). Mean cortical thickness was estimated for preoperative and postoperative images across a set of 74 regions in the Automated Anatomical Labeling (AAL) atlas^31^. Deep brain structures included in the AAL atlas (thalamus, putamen, pallidum, and caudate) were excluded from our analysis as cortical thickness is not estimated for these brain regions. Additionally, regions that overlapped the resection in either treatment group, such as the ipsilateral amygdala, hippocampus, parahippocampus, and temporal pole were excluded from subsequent analyses for both groups. Pre-to-post-operative cortical thinning was calculated by subtracting mean postoperative cortical thickness from mean preoperative cortical thickness of a brain region. To account for potential confounds in the interscan interval (ISI) between treatment groups, annualized cortical thinning was also calculated by dividing the pre-to-post-operative cortical thinning by the ISI. A visual description of the analytical pipeline is provided in **Figure 3**. Patterns of cortical thinning were visualized using the BrainNet Viewer toolbox^32^.

**Figure 1.**
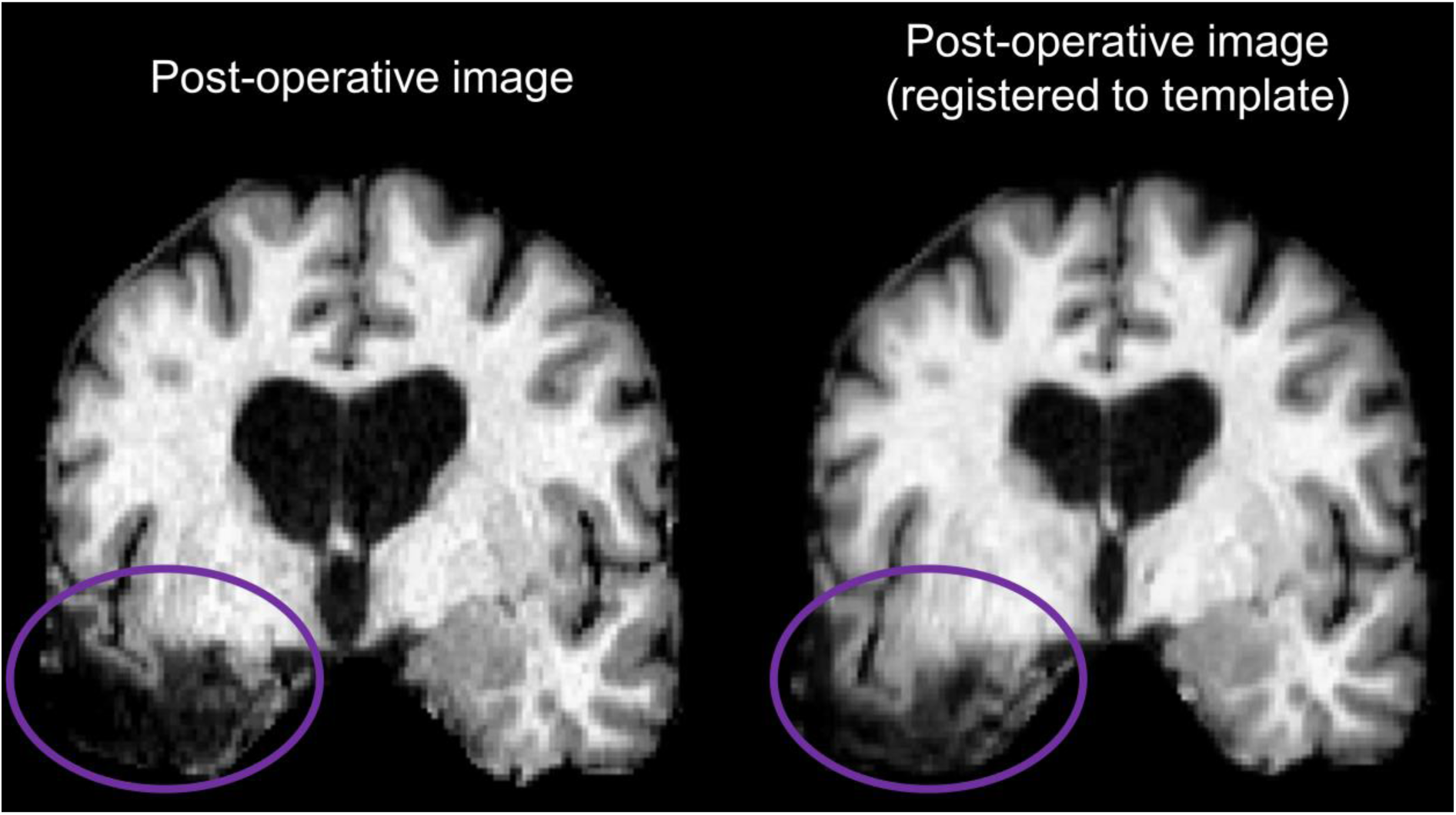
Tissue deformation errors. The original image (left) and the image after registration to a template (right) are shown with purple circles highlighting the area of greatest tissue deformation caused by the registration process.

**Figure 2.**
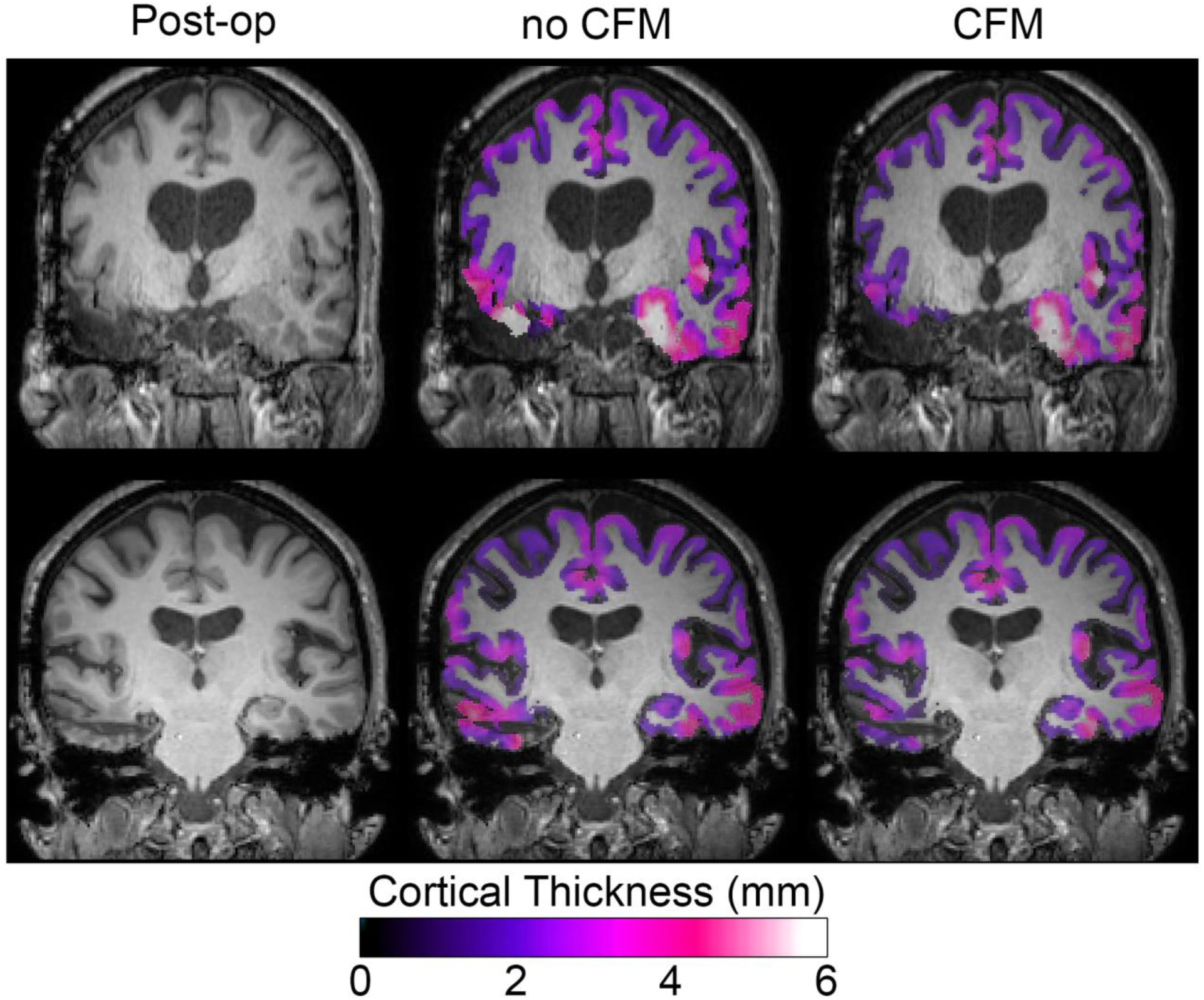
The effect of cost function masking (CFM). The original images (left), cortical thickness estimates without CFM (middle) and cortical thickness estimates with CFM (right) for a subject treat with ATL (top) and another with SAH (bottom) are presented here. Darker colors indicate lower cortical thickness estimates while lighter colors indicate higher cortical thickness values. CFM prevents inaccuracies in cortical thickness estimation near the resected tissue.

**Figure 3.**
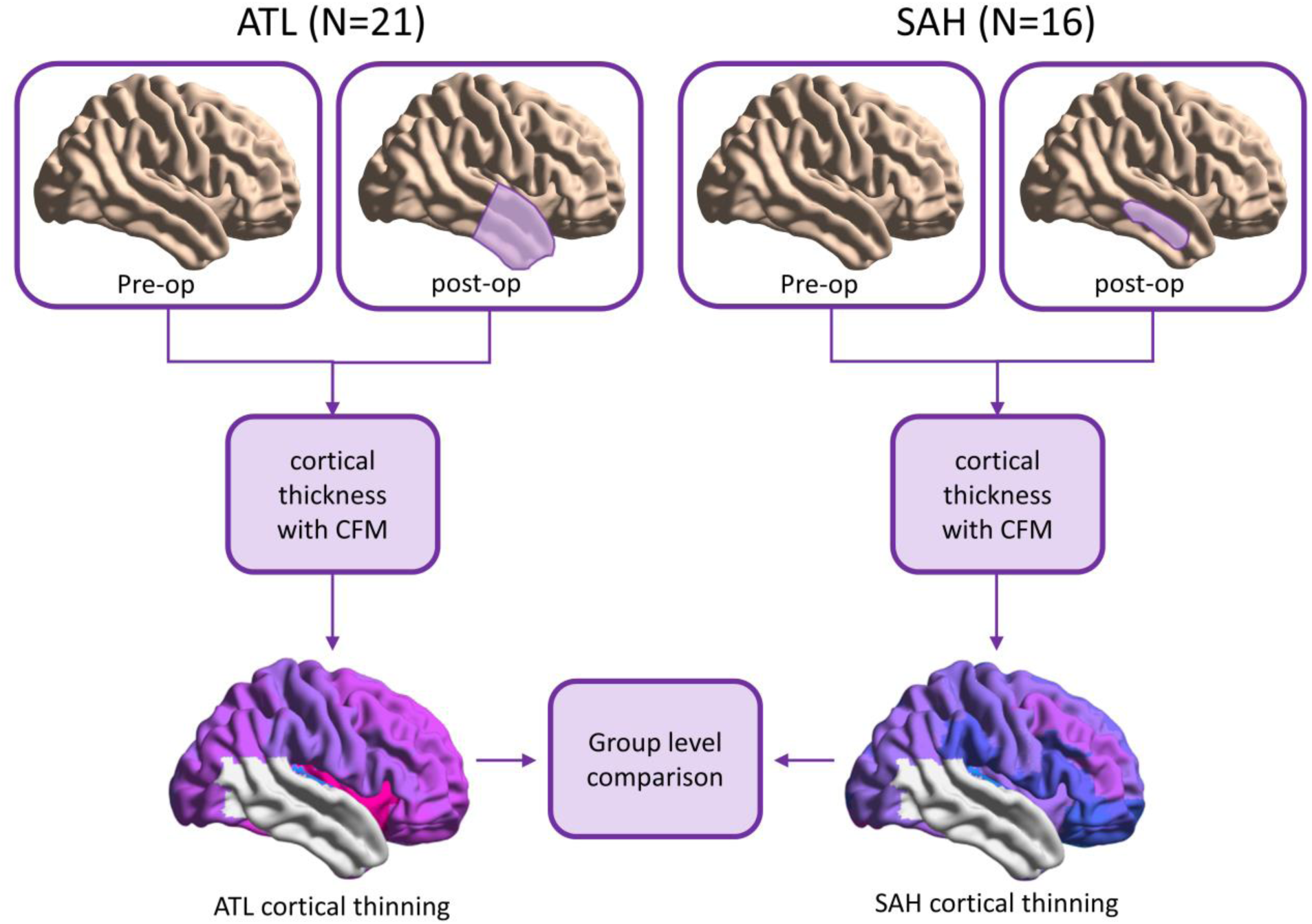
Analytical pipeline. Pre- and post-operative imaging for 21 patients treated with ATL and 14 patients treated with SAH are processed through the ANTs cortical thickness pipeline with a CFM modification. Subsequently each brain region is compared in group level statistical tests. *Abbreviations: ATL, Anterior Temporal Lobectomy; SAH, Selective Amygdalohippocampectomy; CFM, Cost Function Masking; ANT, Advanced Normalization Toolkit*.

### Statistical analysis

Treatment groups were compared for differences in age and gender using a two-sample t-test and chi-squared test respectively. Global differences in pre-to-post-operative cortical thinning were compared between two procedural groups, SAH and ATL, using a two-sample t-test. Differences in cortical thinning were assessed across 74 individual brain regions from the AAL atlas. Brain regions were assessed as either ipsilateral or contralateral to the resection. For all appropriate statistical tests, Bonferroni correction (*α* = 0.05) was applied to account for multiple comparisons. Bonferroni correction across 74 brain regions necessitates a significance threshold of p < 0.00068, which is hereafter referred to as p < 0.001 in statistical reporting by convention.

### Code availability

All code related to resection segmentation, cortical thickness estimation, and statistical analysis are available at: https://github.com/tcama/remote_effects

## RESULTS

### Demographics

Thirty-seven patients were included in this study (14 patients from the Hospital of the University of Pennsylvania and 23 patients from the Vanderbilt University Medical Center). The pipeline failed on one patient due to significant misalignment and segmentation failure as a result of poor imaging quality, and they were excluded from further analysis. Table 1 summarizes important clinical demographics for the study patients.

There was no statistically significant difference between patients treated with SAH or ATL in age (two-sample t-test: t = −0.72, p = 0.48), gender (χ^2^ = 1.05, p = 0.31), or side of seizure onset (χ^2^ = 0.33, p = 0.57). There was a significant difference in the ISI, or time between surgery and postoperative imaging (two-sample t-test: t = 2.91, p = 0.0063), such that patients treated with ATL had a shorter interval (19.4 ± 15.5 month) than those treated with SAH (34.2 ± 15.1 months). Cortical thinning would be expected to be greater for patients with a longer ISI, which could bias analyses towards greater cortical thinning in the SAH treatment group. To address the discrepancy in ISI, we also calculated annualized cortical thinning as explained in the next section.

### Cortical thinning

The degree of postoperative cortical thinning was compared between the two surgical groups across 74 brain regions from the AAL atlas. First, to assess whether there were global differences in cortical thinning between the two-treatment groups, mean cortical thinning was compared using a two-sample t-test (**Figure 4C**). Globally, there was significantly more cortical thinning in patients treated with ATL over those treated with SAH (two-sample t-test: t = 2.04, p = 0.049). This effect was also observed when correcting for differences in the interscan interval using annualized cortical thinning (two-sample t-test: t = 2.86, p = 0.0072). Individually, each group was compared to the null hypothesis using a one-sample t-test. Patients treated with ATL had significantly elevated annualized cortical thinning (0.063 ± 0.095 mm/yr; p = 0.0081), while those treated with SAH did not significantly differ from the null hypothesis (−0.046 ± 0.13 mm/yr; p = 0.19). For all analyses, brain regions overlapping the resection zone in either ATL or SAH patients were excluded from both groups, such that each group contain the same set of regions.

**Figure 4.**
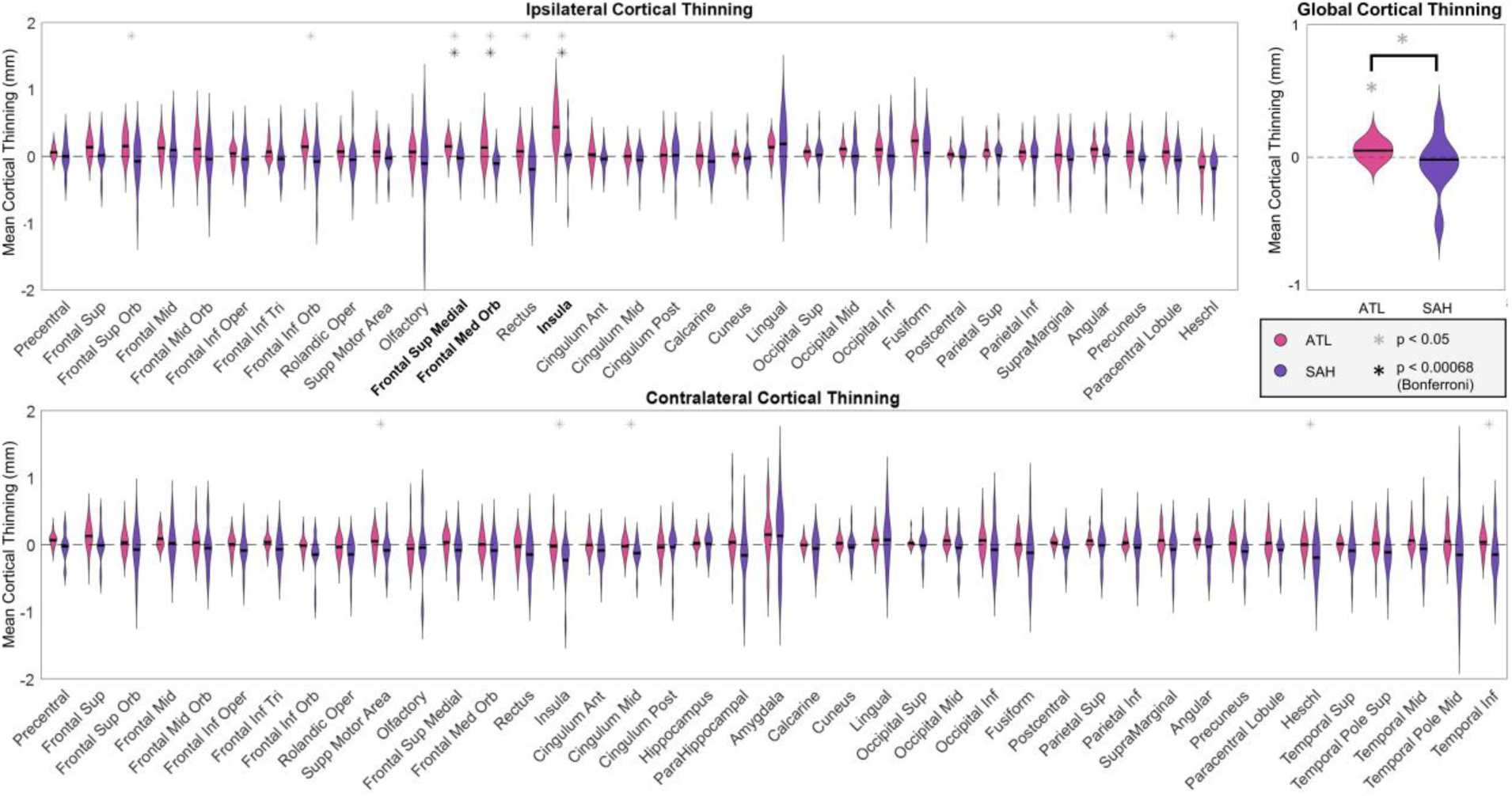
Statistical comparisons of cortical thinning in patients treated with SAH (purple) and ATL (pink). (**A**) Ipsilateral and (**B**) contralateral cortical thinning across 74 brain regions are represented. Note there are more contralateral regions than ipsilateral because the amygdala, hippocampus, parahippocampus, and temporal pole regions were excluded from analysis on the ipsilateral side due to resection overlap. A black asterix indicates significance after Bonferroni correction (p < 0.00068, alpha = 0.05), while a gray asterix indicates uncorrected significance (p < 0.05). The dashed line at zero represents no cortical thinning between the pre- to post-operative time points. The black line on each violin plot represents the median value for the distribution. (**C**) Global cortical thinning was compared to the null hypothesis for each group individually using a one-sample t-test (p <0.05) and between the two groups using a two-sample t-test (p < 0.05). *Abbreviations: ATL, Anterior Temporal Lobectomy; SAH, Selective Amygdalohippocampectomy*.

In the hemisphere ipsilateral to surgery, resections were associated with significantly greater cortical thinning (p < 0.001, *α* = 0.05, Bonferroni corrected for 74 tests) in superior medial (two sample t-test: t = 3.8, p < 0.001) and medial orbital (two sample t-test: t = 3.9, p < 0.001) regions of the frontal lobe as well as the insula (two sample t-test: t = 4.1, p < 0.001, **Figure 4A**) in ATL over SAH. More widespread cortical thinning was observed in the frontal lobe, including the frontal superior orbital, frontal inferior orbital, rectus, and paracentral brain regions. However, these effects did not survive the strict Bonferroni statistical correction applied (p < 0.00068). When accounting for the ISI by comparing the annualized cortical thinning, the effect in the medial orbital region was reduced to a trend-level significance (p = 0.0025, **Figure S1A**), while the insula and superior medial frontal lobe remained significant. In the hemisphere ipsilateral to surgery, most of the effects are driven by an increase in cortical thinning for patients treated with ATL while patients treated with SAH are more likely to remain near baseline (**Figure 4A**).

In the contralateral hemisphere, there were effects in the supplemental motor area, middle cingulum, insula, Heschl’s gyrus, and the inferior temporal lobe, although none of these effects survived rigorous Bonferroni statistical correction (i.e. 0.05 > p > 0.00068, **Figure 4B**). Interestingly, many of the effects in the contralateral hemisphere are driven by *increased cortical thickness* for patients treated with SAH as opposed to *decreased cortical thickness* for ATL patients as observed in the ipsilateral hemisphere. Patients treated with ATL remain near the baseline, while those treated with SAH have negative cortical thinning values, indicating an increase in cortical thickness from pre-to-post-operative imaging.

All effects that were significant before statistical correction are *italicized* in **Table S2** while effects that survived Bonferroni correction listed in **bold**. The extent of cortical thinning in patients treated with ATL and SAH is visualized in **Figure 5A and 5B**, with warmer colors indicating a greater degree of cortical thinning. As observed earlier, there is a global increase in cortical thinning (i.e. warmer colors) for patients treated with ATL in comparisons to those treated with SAH.

**Figure 5.**
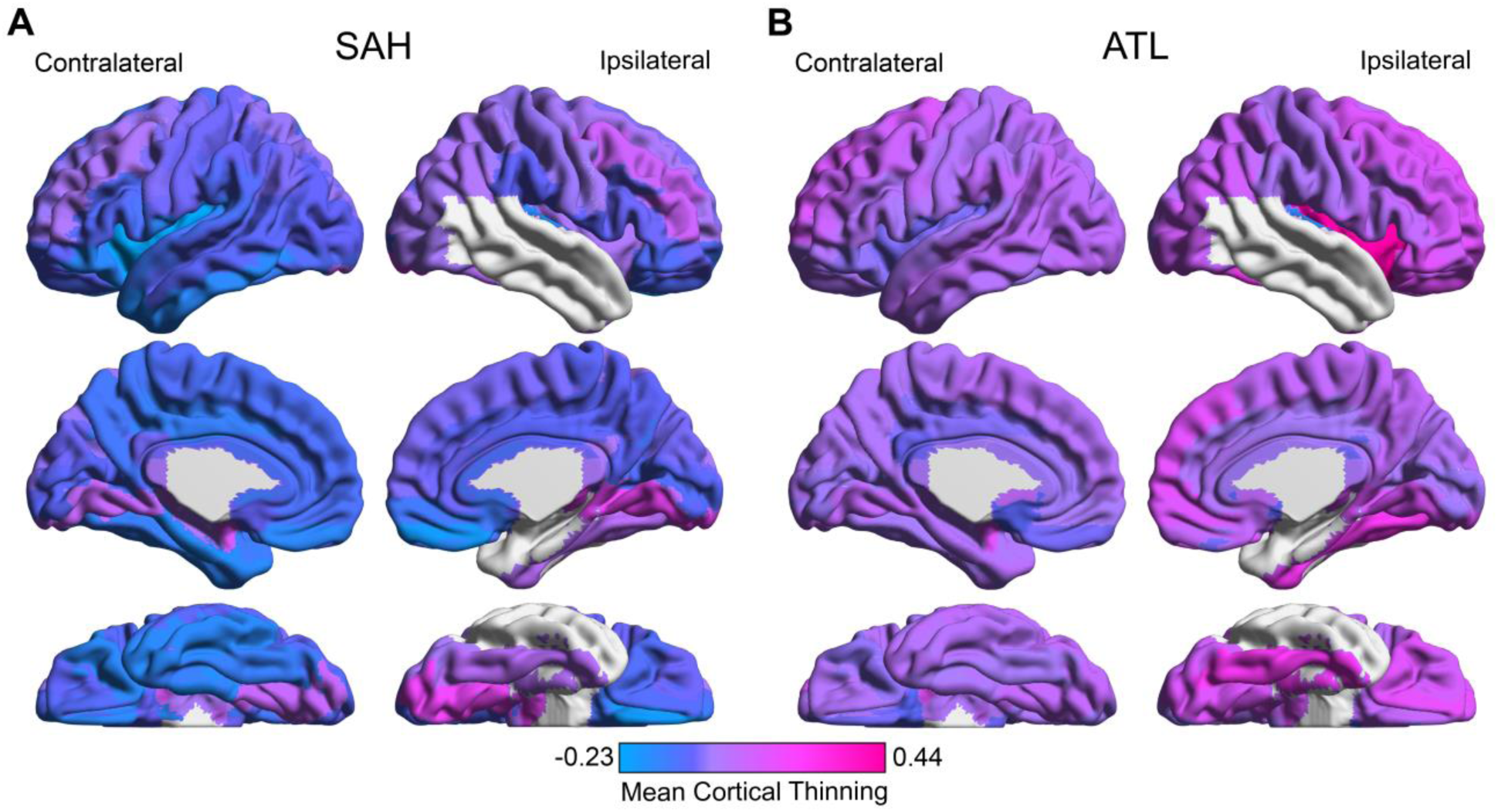
Mean structural changes associated with surgical procedures. The global difference in cortical thinning between the two surgical procedures can be visualized in this comparison of mean cortical thinning patterns. Cooler colors represent areas of low cortical thinning, while hotter colors correspond to high cortical thinning. Overall, there is significantly more cortical thinning globally for patient treated with ATL. *Abbreviations: ATL, Anterior Temporal Lobectomy; SAH, Selective Amygdalohippocampectomy*.

## DISCUSSION

Epilepsy has long been associated with brain atrophy, particularly hippocampal sclerosis in patients with temporal lobe epilepsy^33,34^. Over the past two decades, it has become well established that cortical atrophy in epilepsy is not confined to the epileptogenic zone but rather can be observed throughout the neocortex^35–39^. While the effects of ongoing seizures on cortical thickness have been relatively well studied, comparatively little is known about how surgical operations to treat epilepsy subsequently affect cortical thinning^19,20^. The lack of focus on postsurgical treatment groups in part because of the difficulties in working with postoperative imaging, whereby extra precautions must be taken to ensure that structural alterations do not adversely affect analyses. In this study, we conducted whole-brain comparative analysis of longitudinal cortical thinning between two epilepsy surgical techniques, the anterior temporal lobectomy and the selective amygdalohippocampectomy. To accomplish this goal, we developed semi-automated software for assessing longitudinal cortical thinning while accounting for tissue resected during surgery, which we publicly release.

Our most important finding was that patients treated with SAH had significantly less postoperative cortical thinning than those treated with ATL. While there was a significant effect globally, further analysis revealed that the largest drivers were located in the ipsilateral frontal lobe (superior medial and medial orbital regions) and insula. There were additional effects that did not survive rigorous Bonferroni statistical correction in several ipsilateral and contralateral frontal lobe regions, as well as the contralateral supplemental motor area (SMA), Heschel’s gyrus, and the inferior temporal lobe. Many of the regions in which postoperative benefits of the selective procedure were observed are the same cortices that have been previously associated with progressive cortical thinning due to ongoing epilepsy, including the frontal orbital regions, insula, and Heschl’s gyrus^18,40^.

We speculate that the overlap between preoperative cortical thinning patterns and regional postoperative differences based on surgical approach is reflective of the underlying network connectivity between the affected cortex and downstream regions^41^. Temporal, frontal, and limbic brain regions are known to be integrated into a functional brain network and TLE cortical thinning patterns have been subjectively observed to overlap with regions structural connected to the ipsilateral hippocampus^42,43^. Recent work reveals that epilepsy accelerates cortical atrophy and that regions connected to the epileptogenic zone are most likely to be afflicted with cortical atrophy^42,44^. However, the overlap between preoperative cortical thinning and structural connectivity maps of the putative epileptogenic zone raises the possibility of using cortical thinning as a method for localizing the epileptic focus or differentiating epilepsy subtypes^46^.

Our study adds to a growing body of work that validates cortical thickness measurements as an important biomarker for epilepsy. In a recent longitudinal case-control study, Galovic et al. observed that epilepsy patients had a two-fold increase in the rate of annualized cortical thinning when compared to healthy age-matched controls (TLE 0.024 ± 0.061 mm/yr vs healthy controls 0.011 ± 0.029 mm/yr)^42^. In our own work, we observed that patients treated with ATL had even higher rates of annualized cortical thinning 0.063 ± 0.095 mm/yr (p = 0.0081), while those treated with SAH did not show significant progressive cortical thinning (P = 0.19). Patients treated with ATL demonstrated annualized cortical thinning that was 3 times greater than DRE patients from the Galovic study. The accelerated cortical thinning and differences between the surgical approaches likely reflects greater Wallerian degeneration and loss of functional connections in patients treated with ATL.

One exciting potential for tools that assess postoperative cortical thinning is the possibility of relating quantitative structural analyses and cognitive outcomes. Several studies have compared selective and traditional surgical approaches, with most citing similar seizure outcomes and superior cognitive outcomes for selective procedures^8,9^. However, the cause of these differences in cognitive outcomes remains poorly understood. Selective procedures may preserve more functional gray matter near the focal lesion, or perhaps traditional resections cause greater neurologic injury through trauma to white matter tracts or vasculature, resulting in more widespread effects. In the present study, we demonstrated significant differences in downstream cortical thinning between SAH and ATL. Interestingly, in mesial temporal lobe epilepsy patients Bettus et al. observed a decrease in ipsilateral temporal lobe connectivity while conversely connectivity increased in the contralateral hemisphere^47^. They proposed this may reflect a compensatory mechanism on the contralateral side to preserve normal cognitive function. In our own data, we saw decreased ipsilateral cortical thinning and increased contralateral cortical thickness for patients treated with SAH. This may suggest a structural analog to the compensatory mechanism previously observed. Furthermore, recent work has demonstrated a long term functional connectivity changes in the contralateral hippocampus after surgery^48^. However, the tools outlined in this work would need to be paired with postoperative neuropsychiatric outcomes and functional connectivity to better understand the relationship between postoperative cortical thickness and cognitive outcomes.

Our quantitative cortical thickness methods could provide clinical utility during the surgical planning process, both for epilepsy surgery and other applications. Network-based models provide an exciting *in silico* approach to mapping seizure onset zones and resection zone targeting. New methods, like virtual resection, allow clinicians to map the role of each node using network-control methodology^21,49^. In this case, nodes are representations of electrodes as part of the ECoG or sEEG implant. However, these methods focus solely on the tissues being resected and do not account for downstream effects on the network. The brain regions identified here could be used to modify network-based models to account for such downstream effects. Additionally, recent work by da Silva et al. has demonstrated that epilepsy surgery is associated with changes to structural connectivity in regions remote to the surgical site^50^. By using lesional models of structural connectivity in combination with patterns of postoperative cortical thinning, we can increase our understanding of how focal treatments affect brain structure globally.

Two major limitations of this study are the cohort size and heterogeneity in the ISI. These factors are often limitations in studies of drug resistant epilepsy as obtaining consistent longitudinal data can prove difficult and patients are often lost to follow-up. The present study may be underpowered to detect subtle differences in cortical thinning, especially in subjects with shorter ISI times. While the study may be underpowered, we have still applied a rigorous statistical correction in an attempt to minimize the potential of Type I error. Additionally, annualized cortical thinning analyses were used to address heterogeneity in the ISI. This study represents an initial foray to uncover possible regions of interest that clinicians should note as likely to be affected by resection, however larger studies with consistent ISI should be conducted to establish definitive differences between the treatment groups. Additionally, a comparison to age-matched controls would be desirable, as this would help establish whether patients treated with SAH return to cortical thinning rates similar to the healthy population, which cannot be assessed in the present study. Finally, an additional limitation is the lack of behavioral outcome measures, such as post--resection neuropsychological variables (e.g. memory and language). Future studies will include these outcome variables in addition to new surgical methods such as laser interstitial thermal therapy (LITT), now that our algorithms and methods have been developed and openly shared.

## CONCLUSION

We provide evidence for a relative reduction in postoperative cortical thinning for patients treated with more focused surgical resections. Cortical thinning was globally increased for patients treated with resections, with significant foci in the ipsilateral insula and frontal lobe. Anticipated cortical thinning can inform surgical decisions and postoperative cognitive rehabilitation. In addition, we provide an open-access semi-automated pipeline for the assessment of longitudinal changes in cortical thinning for patients with large structural abnormalities, such as brain tumors or surgical resections.

## Data Availability

The data analyzed in the present study contains sensitive clinical information about patients and must go through formal approval at the collecting institution prior to being shared. It will be made available upon request to the authors, with appropriate institutional data sharing agreements and IRB approval in place. Anonymized data from select subjects are currently publicly available through https://www.ieeg.org/. The code for analyzing postoperative changes in cortical thickness for patients with resections will be made available at https://github.com/tcama/remote_effects. This code relies heavily on the Advanced Normalization Toolkit (ANTs) https://github.com/ANTsX/ANTs. Other code and toolkits are cited appropriately throughout the text. All data processing was done on a Linux server using MATLAB R2019b (The MathWorks Inc., Natick, MA, USA) and Python 2.7 (Python Software Foundation, https://www.python.org/).

## ACKNOWLEDGEMENTS

We would like to acknowledge our sources of support: NIH Virtual cortical resection R01 NS099348 (BL), Pennsylvania Health Research Formula Fund (BL), T32 NS091006 (BL), the Mirowski Family Foundation (BL), The Jonathan Rothberg Family Fund (BL), Neil and Barbara Smit (BL), the Thornton Foundation (KAD), K23 NS073801 (KAD), National Institutes of Health R01 NS075270 (VLM), R01 NS108445 (VLM), R01 NS110130 (VLM), R01 NS112252 (DJE) and R00 NS097618 (DJE) Additional training grant support by HHMI-NIBIB T32 EB009384 Training Program in Biomedical Imaging (TCA).

## ETHICAL PUBLICATION STATEMENT

We confirm that we have read the Journal’s position on issues involved in ethical publication and affirm that this report is consistent with those guidelines.

## SUPPLEMENTAL TABLES & FIGURES

**Table S1 –.** Demographics of patients included in study.

|  | HUP | VUMC | Total |
| --- | --- | --- | --- |
| Number of Participants | 14 | 23 | 37 |
| Gender (Male / Female) | 4/10 | 11/12 | 15/22 |
| Age (Mean ± SD) | 38.9 ± 13.3 | 39.5 ± 12.7 | 39.3 ± 12.7 |
| Seizure side (Left / Right) | 6/8 | 6/17 | 12/25 |
| Surgical Approach (ATL / SAH) | 14/0 | 7/16 | 21/16 |
| Months Since Surgery (Mean ± SD) | 11.3 ± 7.4 | 34.6 ± 14.7 | 25.8 ± 16.9 |

*Note: (1) Months since surgery indicates the interval between the patient's epilepsy surgery and the post-operative imaging. (2) One patient in the VUMC dataset was treated with a Laser Ablation of Left Amygdala & Hippocampus. This patient was included in the SAH group for this study.*
*Abbreviations: HUP, Hospital of the University of Pennsylvania; VUMC, Vanderbilt University Medical Center; SD, Standard Deviation; ATL, Anterior Temporal Lobectomy; SAH, Selective Amygdalohippocampectomy.*

**Table S2 –.** Summary of statistical analysis.

| Ipsilateral regions | p-value |
| --- | --- |
| Frontal (Superior Orbital) | 0.0110 |
| Frontal (Inferior Orbital) | 0.0127 |
| Frontal (Superior Medial) | **0.0006** |
| Frontal (Medial Orbital) | **0.0005** |
| Rectus | 0.0017 |
| Insula | **0.0002** |
| Paracentral lobule | 0.0362 |
| Contralateral regions | p-value |
| Supplementary Motor Area | 0.0405 |
| Olfactory | 0.0218 |
| Cingulum (Middle) | 0.0351 |
| Heschl's gyrus | 0.0055 |
| Temporal (Inferior) | 0.0359 |

*Note: Boldface values indicate statistically significant regions with Bonferroni correction for multiple comparisons. Italic regions represent regions that were statically significant when uncorrected.*

**Figure S1.**
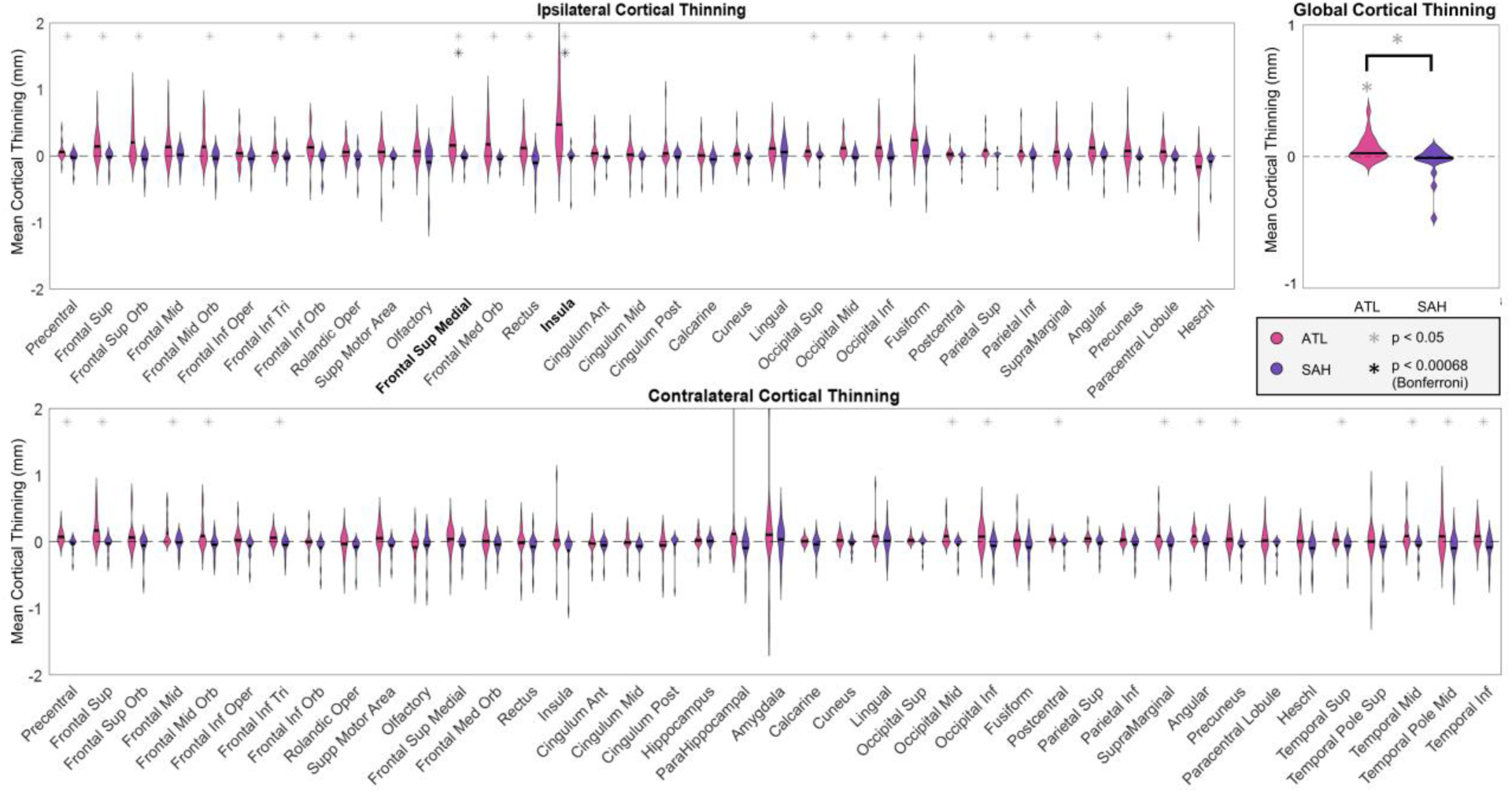
Statistical comparisons of annualized cortical thinning in patients treated with SAH (purple) and ATL (pink). The analysis presented here is identical to the analysis presented in **Figure 4**, except here we present annualized cortical thinning which has been time corrected using the ISI. (**A**) Ipsilateral and (**B**) contralateral annualized cortical thinning across 74 brain regions are represented. Note there are more contralateral regions than ipsilateral because the amygdala, hippocampus, parahippocampus, and temporal pole regions were excluded from analysis on the ipsilateral side due to resection overlap. A black asterix indicates significance after Bonferroni correction (p < 0.00068, alpha = 0.05), while a gray asterix indicates uncorrected significance (p < 0.05). The dashed line at zero represents no cortical thinning between the pre- to post-operative time points. The black line on each violin plot represents the median value for the distribution. (**C**) Global annualized cortical thinning was compared to the null hypothesis for each group individually using a one-sample t-test (p < 0.05) and between the two groups using a two-sample t-test (p < 0.05). *Abbreviations: ATL, Anterior Temporal Lobectomy; SAH, Selective Amygdalohippocampectomy; ISI, InterScan Interval*.

